# Glucagon-like peptide-1 receptor agonists (GLP-1RA) for neuroprotection following aneurysmal subarachnoid haemorrhage (aSAH): a scoping review protocol

**DOI:** 10.1101/2024.12.04.24318321

**Authors:** Matt Thomas, Svetlana Mastitskaya, Sam Parker, Ruth Cookson, Lucy Holmes, Aidan Marsh, Aravind Ramesh, Sarah Rudd, Mario Teo, Alex Mortimer

**Affiliations:** Intensive Care Unit, North Bristol NHS Trust, United Kingdom; Translational Health Sciences, Bristol Medical School, University of Bristol, United Kingdom; Emergency and Critical Care Research Team, North Bristol NHS Trust, United Kingdom; Pharmacy, North Bristol NHS Trust, United Kingdom; Bristol Veterinary School, University of Bristol, United Kingdom; Library and Knowledge Services, North Bristol NHS Trust, United Kingdom; Department of Neurosurgery, North Bristol NHS Trust, United Kingdom; Department of Interventional Neuroradiology, North Bristol NHS Trust, United Kingdom

**Keywords:** cortical spreading depressions, critical care, neurosurgery, stroke, vasospasm, intracranial

## Abstract

**Objective:** The objective of this scoping review is to understand the extent and type of evidence in relation to the use of glucagon-like peptide-1 receptor agonists (GLP1-RA) for neuroprotection in aneurysmal subarachnoid haemorrhage (aSAH).

**Introduction:** The individual and societal costs of aSAH remain high. Effective neuroprotection would reduce morbidity and mortality but there are uncertainties around both established and emerging therapies. GLP1-RA show promise as neuroprotective drugs in other forms of acute and chronic brain injury and could be repurposed for aSAH.

**Inclusion criteria:** Animal and human studies of the use of GLP1-RA for aSAH will be included. Key exclusions are traumatic or non-aneurysmal subarachnoid haemorrhage, or the use of multi-agonist drugs.

**Methods:** Searches will conducted in Embase (Ovid), Medline (Ovid), Cochrane Central Register of Controlled Trials (Wiley) and the World Health Organisation’s International Clinical Trials Registry Platform with no limits applied. Studies for data extraction will be identified by two independent reviewers after screening of titles and abstracts then full text review of potentially relevant articles against inclusion criteria. Data will be extracted by two independent reviewers using a charting tool based on the Joanna Briggs Institute template. A narrative review and summary of findings will be presented.

## Introduction

Stroke is the third most common cause of death globally, and the fourth most common cause of disability adjusted life years. Aneurysmal subarachnoid haemorrhage (aSAH) is the third most common form of stroke, with 0.7 million (95% uncertainty interval 0.5 – 0.8 million) people affected annually worldwide. Overall incidence is 8.3 (7.3 – 9.5) cases per 100,000 population and mortality 4.2 (3.7 – 4.8) per 100,000 with considerable geographic variation.^1^

The decrease in stroke related morbidity and mortality seen from 1990 to 2021 masks a flattening and even an increase of incidence from 2015 onwards, with an increase in the absolute numbers affected by all forms of stroke. Aneurysmal subarachnoid haemorrhage also has a younger peak incidence and increasing prevalence in people under 70 years old.^1^ This, despite reduction in case fatality rates over time,^2^ ^3^ means increasing demand for resources for rehabilitation and greater societal cost of disability with the loss of economically productive individuals. This makes it essential to find effective treatments to improve outcome from aSAH.

Neuroprotection is the preservation, salvage or recovery of central nervous system tissue after acute insult; effective neuroprotective treatments will improve outcomes by reducing death and disability. The current standard neuroprotective treatment for aSAH, nimodipine, is recommended by recent guidelines.^4^ ^5^ Although the assessment of the quality of evidence and the weights of the recommendations differ, there is agreement on the limitations arising from the age of the data and the uncertainties around key outcomes. There are also practical concerns with nimodipine, including hypotension, vasoplegia, a short half life and pharmacokinetic variability, that has led to extensive investigation of alternatives. ^6^

As yet, none of these alternatives are in widespread use in clinical practice.^7^ The lack of effect on long term functional outcomes may reflect a narrow focus on a single relatively late component (arterial vasospasm) of multiple complex pathophysiological processes set in train at ictus. ^8,9^ The opportunity remains for new classes of potentially neuroprotective drugs to be investigated in aSAH.

Glucagon-like peptide-1 receptor agonists (GLP-1RA) are such a class of drug.^10,11^ Actions of GLP-1RA that may be relevant to the pathophysiology of aSAH include modulation of endothelial function, mitigation of ischaemia-reperfusion injury, anti-inflammatory effects, regulation of blood glucose and inhibition of apoptosis, with some evidence of benefit in mice.^12^ In addition they are licensed for other indications in humans, with extensive use demonstrating a low incidence of serious adverse events. However attention to date has largely focused on the use of GLP-1RA for neuroprotection in acute ischaemic stroke and neurodegenerative conditions such as Parkinson’s disease,^13,14^ and whether repurposing for use in aSAH would translate into clinical benefit is as yet unknown.

The aim of this scoping review is to assess the extent of the literature relevant to the use of GLP-1RA for neuroprotection after aneurysmal subarachnoid haemorrhage. A scoping review was chosen because, as noted above, the neuroprotective potential of GLP-1RA has been predominantly investigated in other conditions and current reviews of emerging treatments for aSAH do not include GLP-1RA. ^7,15,16,17^ A preliminary search of PROSPERO, the Cochrane Database of Systematic Reviews, the Open Science Framework and *JBI Evidence Synthesis* was conducted and no current or underway systematic reviews or scoping reviews on the topic were identified.

## Review question

The review question is: “What is the potential role of GLP-1RA in the management of aneurysmal subarachnoid haemorrhage?”

## Inclusion criteria

### Participants

Human and animal studies will be included. All study designs, comparators (including no comparators), and study outcomes will be considered.

### Concept

Studies of aneurysmal subarachnoid haemorrhage will be included. Non-aneurysmal subarachnoid haemorrhage, including traumatic and perimesencephalic subarachnoid haemorrhage, will be excluded.

Glucagon-like peptide-1 receptor agonists, licensed, unlicensed and experimental will be considered. Multi-agonists will be excluded.

### Context

Aneurysmal subarachnoid haemorrhage is primarily a disease of adults and is often managed in Intensive Care Units; these will not however be restrictions applied to this review. There will be no restrictions based on culture, location or any protected characteristic.

## Types of sources

This scoping review will consider both experimental and quasi-experimental study designs in humans and animals including randomized controlled trials, non-randomized controlled trials, before and after studies and interrupted time-series studies. In addition, analytical observational studies including prospective and retrospective cohort studies, case-control studies and analytical cross-sectional studies will be considered for inclusion. This review will also consider descriptive observational study designs including case series, individual case reports and descriptive cross-sectional studies for inclusion. The reference lists of all articles retrieved for full text analysis as well as the authors’ personal collections will be searched for additional information sources.

## Methods

The proposed scoping review will be conducted in accordance with the JBI methodology for scoping reviews.^18^ The review was registered prospectively on 4th October 2024 on the Open Science Framework (osf.io/v95d7).

### Search strategy

A systematic search of four electronic bibliographic databases (Embase (Ovid), Medline (Ovid), Cochrane Central Register of Controlled Trials (Wiley) and the World Health Organisation’s International Clinical Trials Registry Platform (ICTRP)) will be conducted in line with Cochrane Handbook ^19^and JBI Manual for Evidence Synthesis^18^ recommendations and other systematic reviews investigating the use of GLP-1 receptor agonists.^20,21^ ^22^ The search strategy will include keyword and subject heading search terms relating to subarachnoid haemorrhage and GLP-1RA’s (including exenatide or liraglutide or albiglutide or dulagutide or lixisenatide or semaglutide or tirzepatide). No limits or filters will be applied to the searches.

### Study/Source of evidence selection

Following the search, all identified citations will be collated and uploaded into Rayyan (https://www.rayyan.ai/) and duplicates removed. Following a pilot test, titles and abstracts will then be screened by two independent reviewers for assessment against the inclusion criteria for the review, with discrepancies adjudicated by a third reviewer. Potentially relevant sources will be retrieved in full and their citation details imported into the JBI System for the Unified Management, Assessment and Review of Information (JBI SUMARI) (JBI, Adelaide, Australia).^23^

The full text of selected citations will be assessed in detail against the inclusion criteria by two independent reviewers. Reasons for exclusion of sources of evidence after full text review will be recorded and reported in the scoping review. Disagreements that arise between the reviewers will be resolved by a third reviewer. The results of the search and the study inclusion process will be reported in full in the final scoping review and presented in a PRISMA flow diagram.^24^

### Data extraction

Data will be extracted from papers included in the scoping review by two independent reviewers using a data extraction tool developed by the reviewers from the JBI data charting template.^18^ The data extracted will include specific details about the participants, concept, context, study methods and key findings relevant to the review question/s. Critical appraisal of the evidence will not be undertaken.

A draft extraction form is provided (see Appendix II). The draft data extraction tool will be modified and revised as necessary during the process of extracting data from each included evidence source. Modifications will be detailed in the scoping review. Any disagreements that arise will be resolved by a third reviewer. Where discrepancies are identified that cannot be resolved by the third reviewer the original investigators will be contacted by email with a follow up after two weeks if no response is not received.

### Data analysis and presentation

Evidence will be presented as narrative summaries categorised according to species (human/animal) and as summary tables. The evidence will not be critically appraised or subject to statistical analysis (including meta-analysis).

## Data Availability

All data that will be produced in the work described in this protocol will be contained in the review manuscript

## Acknowledgements

No acknowledgements.

## Declarations

No declarations.

## Author contributions

AMa - data collection, manuscript writing and review

AMo - concept, study design, manuscript writing and review

AR - manuscript review

LH - manuscript review

MT - concept, study design, data collection, manuscript writing and review

RC - manuscript review

SM - study design, manuscript writing and review

SR - literature search, manuscript review

SP - data collection, manuscript writing and review

## Appendices

### Appendix I: Search strategy

Search Strategies

1. Embase (Ovid)

1. subarachnoid hemorrhage/
2. exp brain hemorrhage/
3. exp carotid artery aneurysm/
4. exp brain hematoma/
5. ((subarachnoid* or arachnoid* or brain* or cerebral or intracerebral or intracranial or cerebell* or intraventricular or parenchymal or infratentorial or supratentorial or carotid) adj4 (h?errhag* or h?ematoma or bleed* or blood or vasospasm* or vasoconstrict* or angiospasm*)).ti,ab,kf.
6. exp brain vasospasm/
7. ”delayed cerebral isch?emia”.ti,ab,kf.
8. ((cortical or cortex or spreading) adj3 (depolarisation or depression)).ti,ab,kf.
9. spreading cortical depression/
10. 1 or 2 or 3 or 4 or 5 or 6 or 7 or 8 or 9
11. (exenatide or “AC 2993” or AC2993 or “ITCA 650” or ITCA650 or “exendin 4”).ti,ab,kf.
12. exendin 4/
13. 13 exenatide.dy.
14. exendin 4.dy.
15. 15 liraglutide/
16. (liraglutide or “NN 2211” or NN2211 or “NNC 90 1170” or “NNC90 1170”).ti,ab,kf.
17. 17 liraglutide.dy.
18. 18 albiglutide/
19. (albiglutide or GSK 716155 or GSK716155 or (albumin adj3 (”GLP 1” or GLP1 or “glucagon like peptide”))).ti,ab,kf.
20. 20 albiglutide.dy.
21. 21 dulaglutide/
22. (dulaglutide or LY2189265 or “LY 2189265”).ti,ab,kf.
23. 23 dulaglutide.dy.
24. 24 lixisenatide/
25. (lixisenatide or “AVE 0010” or AVE0010).ti,ab,kf.
26. 26 lixisenatide.dy.
27. 27 semaglutide.dy.
28. 28 semaglutide/
29. (semaglutide or “NN 9535” or NN9535).ti,ab,kf.
30. 30 tirzepatide/
31. 31 Tirzepatide.dy.
32. (Tirzepatide or “LY-3298176” or LY3298176 or Mounjaro).ti,ab,kf.
33. ((glucagon like peptide* or “GLP 1” or GLP1) adj3 (analog* or agonist*)).ti,ab,kf.
34. ”Incretin mimeti*”.ti,ab,kf.
35. (”GLP1-RA” or GLP1RA or “GLP-1-RA”).ti,ab,kf.
36. glucagon like peptide 1 receptor agonist/
37. 37 or/11-36
38. 10 and 37

2. Medline All (Ovid)

Ovid MEDLINE(R) ALL <1946 to MMM DD, 2024>

1. exp Subarachnoid Hemorrhage/
2. exp Intracranial Hemorrhages/
3. Intracranial Aneurysm/
4. exp Hematoma, Subdural/
5. ((subarachnoid* or arachnoid* or brain* or cerebral or intracerebral or intracranial or cerebell* or intraventricular or parenchymal or infratentorial or supratentorial or carotid) adj4 (h?errhag* or h?ematoma or bleed* or blood or vasospasm* or vasoconstrict* or angiospasm*)).ti,ab,kf.
6. Vasospasm, Intracranial/
7. ”delayed cerebral isch?emia”.ti,ab,kf.
8. ((cortical or cortex or spreading) adj3 (depolarisation or depression)).ti,ab,kf.
9. Cortical Spreading Depression/
10. 1 or 2 or 3 or 4 or 5 or 6 or 7 or 8 or 9
11. (exenatide or “AC 2993” or AC2993 or “ITCA 650” or ITCA650 or “exendin 4”).ti,ab,kf.
12. Exenatide/
13. Liraglutide/
14. (liraglutide or “NN 2211” or NN2211 or “NNC 90 1170” or “NNC90 1170”).ti,ab,kf.
15. (albiglutide or GSK 716155 or GSK716155 or (albumin adj3 (”GLP 1” or GLP1 or “glucagon like peptide”))).ti,ab,kf.
16. (dulaglutide or LY2189265 or “LY 2189265”).ti,ab,kf.
17. (lixisenatide or “AVE 0010” or AVE0010).ti,ab,kf.
18. (semaglutide or “NN 9535” or NN9535).ti,ab,kf.
19. (Tirzepatide or “LY-3298176” or LY3298176 or Mounjaro).ti,ab,kf.
20. ((glucagon like peptide* or “GLP 1” or GLP1) adj3 (analog* or agonist*)).ti,ab,kf.
21. ”Incretin mimeti*”.ti,ab,kf.
22. (”GLP1-RA” or GLP1RA or “GLP-1-RA”).ti,ab,kf.
23. Glucagon-Like Peptide-1 Receptor Agonists/
24. 11 or 12 or 13 or 14 or 15 or 16 or 17 or 18 or 19 or 20 or 21 or 22 or 23
25. 10 and 24

• 3. Cochrane CENTRAL (Wiley)

ID Search Hits

#1 MeSH descriptor: [Subarachnoid Hemorrhage] explode all trees

#2 MeSH descriptor: [Intracranial Hemorrhages] explode all trees

#3 MeSH descriptor: [Intracranial Aneurysm] explode all trees

#4 MeSH descriptor: [Hematoma, Subdural] explode all trees

#5 (((subarachnoid* or arachnoid* or brain* or cerebral or intracerebral or intracranial or cerebell* or intraventricular or parenchymal or infratentorial or supratentorial or carotid) Near/4 (h?errhag* or h?ematoma or bleed* or blood or vasospasm* or vasoconstrict* or angiospasm*))):ti,ab,kw

#6 MeSH descriptor: [Vasospasm, Intracranial] explode all trees

#7 “delayed cerebral” NEXT isch?emia

#8 (((cortical or cortex or spreading) NEAR/3 (depolarisation or depression))):ti,ab,kw

#9 MeSH descriptor: [Cortical Spreading Depression] explode all trees

#10 #1 or #2 or #3 or #4 or #5 or #6 or #7 or #8 or #9

#11 ((exenatide or “AC 2993” or AC2993 or “ITCA 650” or ITCA650 or “exendin 4”)):ti,ab,kw

#12 MeSH descriptor: [Exenatide] explode all trees

#13 MeSH descriptor: [Liraglutide] explode all trees

#14 ((liraglutide or “NN 2211” or NN2211 or “NNC 90 1170” or “NNC90 1170”)):ti,ab,kw

#15 ((albiglutide or “GSK 716155” or GSK716155 or (albumin NEAR/3 (”GLP 1” or GLP1 or “glucagon like peptide”)))):ti,ab,kw

#16 ((dulaglutide or LY2189265 or “LY 2189265”)):ti,ab,kw

#17 ((lixisenatide or “AVE 0010” or AVE0010)):ti,ab,kw

#18 ((semaglutide or “NN 9535” or NN9535)):ti,ab,kw

#19 ((Tirzepatide or “LY-3298176” or LY3298176 or Mounjaro)):ti,ab,kw

#20 (((glucagon like peptide* or “GLP 1” or GLP1) NEAR/3 (analog* or agonist*))):ti,ab,kw

#21 (Incretin NEXT mimeti*):ti,ab,kw

#22 ((”GLP1-RA” or GLP1RA or “GLP-1-RA”)):ti,ab,kw

#23 MeSH descriptor: [Glucagon-Like Peptide-1 Receptor Agonists] explode all trees

#24 #11 or #12 or #13 or #14 or #15 or #16 or #17 or #18 or #19 or #20 or #21 or #22 or #23

#25 #10 and #24

4. WHO ICTRP

1. (hemorrhag* or haemorrhage or hematom* or haematom*) AND (exenatide or liraglutide or albiglutide or dulagutide or lixisenatide or semaglutide or tirzepatide)
2. (exenatide or liraglutide or albiglutide or dulagutide or lixisenatide or semaglutide or tirzepatide) and (subarachnoid* or arachnoid* or brain* or cerebral or intracerebral or intracranial or cerebell* or intraventricular or parenchymal or infratentorial or supratentorial or carotid)
3. (”glucagon like peptide*” or “GLP 1” or GLP1) and (subarachnoid* or arachnoid* or brain* or cerebral or intracerebral or intracranial or cerebell* or intraventricular or parenchymal or infratentorial or supratentorial or carotid)

### Appendix II: Data extraction instrument

Data charting tool v1.0

Paper for inclusion reference number

Evidence source details/characteristics

Citation

Species

Setting

Clinical / laboratory

Type of evidence source

Study design/characteristics

Design

Participants (numbers, age, sex etc)

Intervention

Comparator

Outcome(s)

Results extracted

Outcome (describe) Data

Outcome (describe) Data

Outcome (describe) Data

Outcome (describe) Data

Outcome (describe) Data

